# Who is and who is not attempting to quit smoking? A population study in Great Britain, 2020–2026

**DOI:** 10.64898/2026.08.05.26359774

**Authors:** Sarah E. Jackson, Deborah Robson, Jamie Brown, Caitlin Notley, Claire Garnett, Sharon Cox

## Abstract

**Aims:** To estimate the prevalence of past-year smoking quit attempts in Great Britain and examine variation by sociodemographic and socioeconomic characteristics, mental health, alcohol use, smoking-related characteristics, and geographic area.

**Design:** Cross-sectional analysis of data from a nationally representative household survey (the Smoking Toolkit Study) conducted October 2020 to May 2026.

**Setting:** Great Britain.

**Participants:** 24,786 adults (≥18y) who reported past-year tobacco smoking.

**Measurements:** The outcome was self-reporting having made at least one serious attempt to stop smoking in the previous 12 months. Associations with age, gender, ethnicity, social grade, health-related economic inactivity, mental health conditions, psychological distress, alcohol consumption, use of non-combustible nicotine, cigarette type and consumption, strength of urges to smoke, and motivation to stop smoking were assessed. We calculated weighted prevalence estimates and odds ratios (ORs) adjusted for survey year.

**Findings:** Overall, 36.7% [95%CI=36.0-37.4] of adults who had smoked in the past year self-reported making at least one quit attempt. Annual prevalence was relatively stable over the study period (range: 35.5% [33.9-37.1] to 37.6% [35.9-39.3]). Making a past-year quit attempt was more common among younger adults (48.0% among 18-24-year-olds) and declined progressively with age (24.6% among ≥65-year-olds; OR=0.35, 95%CI=0.31-0.39). Compared with White adults, making a past-year quit attempt was more common among Black (OR=1.31, 1.11-1.55) and Asian (OR=1.40, 1.21-1.62) adults. Adults with a history of diagnosed mental health conditions (OR=1.27, 1.17-1.38) and moderate (OR=1.31, 1.20-1.43) or severe psychological distress in the past month (OR=1.39, 1.25-1.56) had greater odds of reporting a past-year quit attempt than those without. Those reporting increasing/higher risk alcohol consumption had lower odds than non-drinkers (OR=0.88, 0.82-0.95). Current use of nicotine replacement therapy (OR=2.59, 2.36-2.83) and vapes (OR=2.15, 2.02-2.30) were positively associated with reporting a past-year quit attempt. Odds were lower among those with greater cigarette consumption (OR range 0.79-0.86 among those smoking >10 vs. ≤5 cigarettes per day). Motivation to stop smoking showed the strongest gradient (OR range 2.95-29.09). Geographic differences were modest, with prevalence ranging from 32.1% [29.9-34.3] in Wales to 39.4% [35.9-43.1] in North East England.

**Conclusions:** Between 2020 and 2026, around one in three adults in Great Britain who smoked in the past year reported making a serious attempt to quit, corresponding to approximately 3.5 million people annually. Making a past-year quit attempt was more strongly associated with smoking-related factors than sociodemographic characteristics.

## Introduction

Smoking remains a leading preventable cause of disease and premature death in Great Britain^1^ despite substantial progress in tobacco control over recent decades.^2,3^ The recent passage of the UK Tobacco and Vapes Act 2026,^4^ including the introduction of a ‘smokefree generation’ policy from January 2027 whereby anyone born after 2008 can never legally be sold tobacco, is a key step in preventive health policy. However, while this policy is designed to prevent initiation among future cohorts, it does not address the needs of more than five million adults in the UK who continue to smoke,^3^ among whom major inequalities persist.^5–9^

Two key mechanisms for achieving population-level declines in smoking prevalence are increasing the rate of quit attempts and increasing the likelihood that quit attempts are successful. In recognition of this, the government has recently increased investment in stop smoking services provided to local government and the NHS, including by £70 million a year^10^ for local government – more than doubling previous spending^5^ – and £35m in 2022 to the NHS,^10^ with the aim of expanding access to behavioural and pharmacological support. Early evidence suggests that service throughput is increasing,^11^ and these developments are expected to improve quit success among those who engage with support services.^12,13^ However, sustained reductions in smoking prevalence will also depend on encouraging more people who smoke to make quit attempts.

Evidence consistently shows that the majority of adults who smoke do not make a quit attempt in a given year,^14–18^ and prior research has examined predictors of quit attempts in general population samples.^16,19–21^ These studies suggest that motivational factors are the most consistent correlates of making a quit attempt, including intention to quit, previous quit attempts, and cognitive evaluations of smoking such as concern about health and perceived benefits of stopping.^16,19,20^ Indicators of nicotine dependence (e.g., cigarettes per day and dependence scores) are also frequently associated with lower likelihood of attempting to quit,^19,20^ although findings are not entirely consistent across studies.^19^ In contrast, sociodemographic characteristics (including age, gender, and socioeconomic position) show generally weak or inconsistent associations with quit attempts across studies.^16,19–24^

However, much of this evidence predates recent changes in the tobacco control and wider social context, including the COVID-19 pandemic,^15^ the cost-of-living crisis,^25^ and expansions in cessation support provision. It also predates recent policy developments in Great Britain aimed at accelerating progress towards a smokefree future,^4,10^ alongside smoking prevalence reaching historic lows.^3^ Updated evidence is therefore needed to identify which groups are currently less likely to try to quit smoking, to support the design, targeting, and delivery of contextually relevant interventions, including public health campaigns and cessation services, in the context of forthcoming new policies.

The Smoking Toolkit Study provides an opportunity to examine patterns of smoking quit attempts among adults in Great Britain using a large, nationally representative dataset with detailed information on sociodemographic characteristics, smoking behaviour, mental health, and alcohol use. Using data from this survey, this study aimed to estimate the prevalence of past-year smoking quit attempts among adults who smoke, both overall and within key subgroups defined by sociodemographic and socioeconomic characteristics, mental health, alcohol use, non-combustible nicotine use, smoking-related characteristics, and geographic area. These characteristics were selected because they are recognised determinants of smoking and cessation behaviour and may identify groups experiencing inequalities in quitting or requiring targeted cessation support. In addition, we compared the profile of all adults who smoked in the past year, those who made a quit attempt, and those who did not, across these domains.

## Methods

### Pre-registration

The study protocol and analysis plan were pre-registered on Open Science Framework (https://osf.io/gw4mn). In addition to our planned analyses, we examined associations between motivation to stop smoking and quit attempts among those who currently smoked, to provide a more complete picture of factors associated with quit attempts. We also conducted analyses adjusted for age, gender, ethnicity, and social grade to assess the extent to which associations between quit attempts and other characteristics were confounded by sociodemographic factors.

### Design

The Smoking Toolkit Study is an ongoing monthly cross-sectional survey of a representative sample of adults (≥16 years) in Great Britain.^26,27^ Each month, approximately 2,450 adults are recruited using a hybrid sampling approach combining random probability and simple quota sampling. Self-reported data are collected by market research company Ipsos via telephone interviews. The study has been shown to produce nationally representative estimates of key indicators, including sociodemographic characteristics, smoking prevalence, and cigarette consumption, when compared with external benchmarks such as national surveys and sales data.^26,28^

The present analyses used data collected between October 2020 (the first data collection in Wales and Scotland) and May 2026 (the most recent data available at the time of analysis). The sample was restricted to respondents who reported that they currently smoked tobacco (daily or non-daily) or had stopped smoking completely in the past year (i.e., all adults who had smoked in the past year). Because 16–17-year-olds were not included in the survey between October 2020 and December 2021, the analytic sample was further restricted to adults aged ≥18 years to ensure consistency across the study period. We excluded participants with missing data on past-year quit attempts.

### Measures

Full details of the measures are provided in the study protocol (https://osf.io/gw4mn).

Past-year quit attempts were assessed with the question: ‘How many serious attempts to stop smoking have you made in the last 12 months?’. A serious attempt was defined as deciding to stop smoking permanently and included ongoing and successful attempts made within the past year. Participants reporting one or more attempts were classified as having made a quit attempt.

Sociodemographic characteristics included age group, gender, ethnicity, and occupational social grade (based on National Readership Survey classifications^29^). Health-related economic inactivity was classified as reporting not being in paid work because of long-term illness or disability. Geographic area was categorised based on nation (England, Wales, or Scotland) and the nine regions of England.

Mental health variables included diagnosed mental health conditions and past-month psychological distress. We derived a binary (yes/no) variable reflecting ever being diagnosed with any of the following mental health conditions since the age of 16: depression; anxiety; bipolar disorder; obsessive compulsive disorder; panic disorder or a phobia; post-traumatic stress disorder; psychosis; personality disorder; an eating disorder; alcohol misuse or dependence; drug use disorder, drug abuse or drug dependence; or problem gambling.

Psychological distress was assessed using the Kessler Psychological Distress Scale (K6), categorised as no/low distress (scores of 0-4), moderate distress (5-12), or severe distress (13-24).^30–32^ These two variables were assessed in a subset of waves (October 2020-June 2023, February, April, June, August, October, and December 2025, plus January-March 2024 for distress only), so relevant analyses were restricted to participants surveyed in these months. These two variables were assessed among all participants in England during these waves and all participants in Wales and Scotland in the 2025 waves, but only among approximately 50% of participants in Wales and Scotland in the 2020-2024 waves, due to funding limitations.

Alcohol consumption was assessed with the Alcohol Use Disorders Identification Test-Consumption (AUDIT-C) and categorised as non-drinking (scores of 0), low risk (1-4), or increasing/higher risk (5-12).^33,34^ We also included measures of non-combustible nicotine, including current use vs. non-use of nicotine replacement therapy (NRT), vapes, heated tobacco products, and nicotine pouches.

Smoking-related characteristics included strength of urges to smoke over the last 24 hours,^37^ assessed among all participants who reported smoking in the past year, daily cigarette consumption (‘How many cigarettes do/did you usually smoke?’)^35^ and the main type of cigarettes (manufactured or hand-rolled)^36^ among those who smoked cigarettes in the past year, and motivation to stop smoking^38^ among those who reported current smoking.

Survey year was analysed as a categorical variable from 2020 to 2026. Note that 2020 includes data from October-December only and 2026 from January-May only.

### Statistical analysis

Analyses were conducted in R v.4.6.0. The Smoking Toolkit Study applies raking weights to ensure the sample is representative of the adult population in Great Britain. This profile is determined each month by combining data from the UK Census, the Office for National Statistics mid-year estimates, and the annual National Readership Survey.^39^ All analyses used weighted data to provide population-representative estimates, with the exception of analyses stratified by region in England, which used England-specific weights. Analyses involving mental health conditions and psychological distress used different weights to account for the reduced subsample in which these measures were collected between 2020 and 2024. Sample sizes are reported unweighted.

Missing data were handled using complete-case analysis on a per-analysis basis. The exception was social grade, where missing cases (<5%) were recoded as C1 in accordance with the standard Smoking Toolkit Study weighting protocol, which assumes missing values are C1 when deriving the weight variable, ensuring consistency between weighted analyses and analyses including social grade.

We described the characteristics of: (1) all adults who smoked in the past year, (2) those who made at least one quit attempt, and (3) those who did not attempt to quit. For each group, we reported proportions (with 95% confidence intervals [CI]) across sociodemographic, mental health, alcohol use, non-combustible nicotine use, and all smoking-related variables.

Among adults who smoked in the past year, we estimated the proportion (with 95% CI) who reported making at least one past-year quit attempt and the proportion who did not make a quit attempt, both overall and by survey year. To estimate the annual number of adults in Great Britain making smoking quit attempts, we combined mid-year population estimates from the Office for National Statistics^40^ with annual estimates of past-year smoking prevalence from the Smoking Toolkit Study. The annual number of quit attempts was estimated as:

*total number of adults in Great Britian × proportion who smoked in the past year × proportion of those who smoked in the past year who made a past — year smoking quit attempt.*

We also estimated the prevalence of having made at least one past-year smoking quit attempt stratified by sociodemographic, mental health, alcohol use, non-combustible nicotine use, and smoking-related variables. Geographic variation was visualised using heatmaps.

We used a series of logistic regression models to examine associations between each of these participant characteristics (excluding region in England) and having made a past-year quit attempt (yes vs. no). Each association was tested in a separate model, and models were adjusted for survey year to account for any temporal variation across the study period. We assessed whether associations between participant characteristics and quit attempts varied over time by repeating these models including interaction terms between each variable and survey year.

In unplanned analyses, we repeated the models (without interaction terms) with additional adjustment for age, gender, ethnicity, and social grade to explore the extent to which associations with other variables were explained by sociodemographic factors. We also repeated the models testing associations with non-combustible nicotine use, cigarette consumption and type, and strength of urges to smoke among those who reported current smoking only, to examine whether estimates differed when excluding participants who had successfully quit within the past year and whose smoking-related characteristics may not reflect their status at the time of quitting.

## Results

A total of 155,867 adults aged ≥18 years were surveyed in Great Britain between October 2020 and May 2026, of whom 25,835 reported current smoking or having quit in the past year (i.e., past-year smoking). We excluded 1,049 participants (4.1%) with missing data on past-year quit attempts, resulting in a final analytic sample of 24,786 participants. Numbers of missing cases on other variables are reported in **Table S1**.

Sample characteristics are presented in **Tables 1-2**, overall and stratified by past-year smoking quit attempts. Associations of past-year quit attempts with sociodemographic, mental health, alcohol use, non-combustible nicotine use, and smoking-related characteristics are shown in **Tables 3-4**. A narrative comparison of the profile of those who made a quit attempt vs. those who did not is provided in the **Supplementary File**.

**Table 1.**
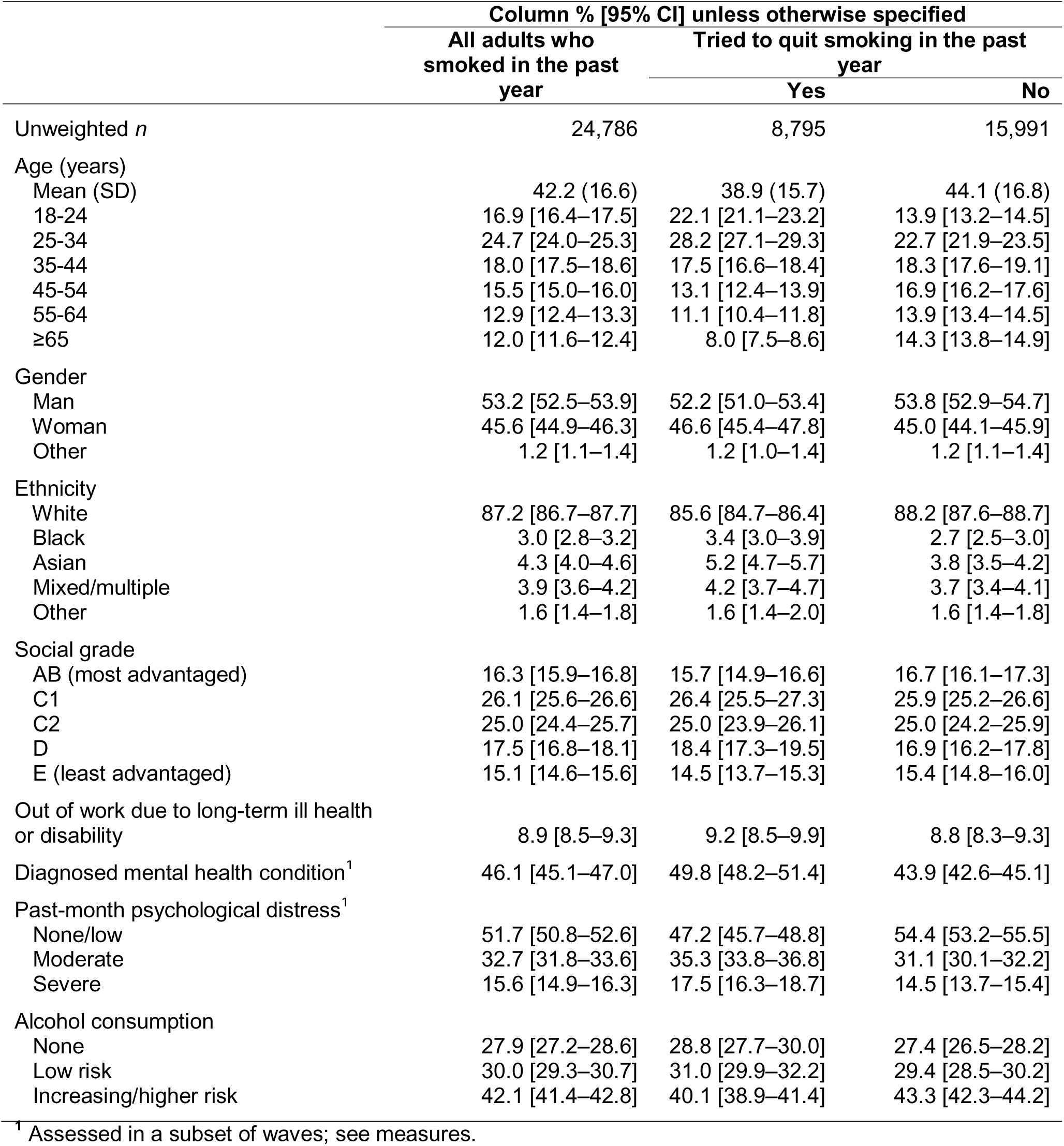
Sociodemographic, socioeconomic, mental health, and alcohol use profile of those who did and did not make a past-year smoking quit attempt.

**Table 2.** Non-combustible nicotine use and smoking profile of those who did and did not make a past-year smoking quit attempt.

|  | Column % [95% CI] |  |  |
| --- | --- | --- | --- |
|  | All adults who smoked in the past year | Tried to quit smoking in the past year |  |
|  |  | Yes | No |
| Unweighted *n* | 24,786 | 8,795 | 15,991 |
| Current use of... |  |  |  |
| Nicotine replacement therapy | 11.3 [10.8–11.7] | 17.6 [16.7–18.5] | 7.6 [7.2–8.1] |
| Vapes | 30.1 [29.4–30.8] | 40.4 [39.2–41.6] | 24.2 [23.4–24.9] |
| Heated tobacco products | 1.0 [0.9–1.2] | 1.3 [1.0–1.6] | 0.9 [0.7–1.0] |
| Nicotine pouches | 2.4 [2.2–2.7] | 3.3 [2.9–3.8] | 1.9 [1.7–2.2] |
| Daily cigarette consumption<sup>1</sup> |  |  |  |
| Mean (SD) | 10.6 (9.0) | 10.3 (8.7) | 10.8 (9.1) |
| ≤5 | 35.0 [34.3–35.8] | 37.1 [35.9–38.3] | 33.7 [32.8–34.7] |
| 6-10 | 28.9 [28.2–29.6] | 28.9 [27.8–30.1] | 28.9 [28.0–29.8] |
| 11-15 | 15.3 [14.8–15.9] | 14.1 [13.3–15.0] | 16.1 [15.4–16.8] |
| 16-20 | 14.8 [14.3–15.4] | 14.3 [13.4–15.2] | 15.1 [14.5–15.9] |
| >20 | 5.9 [5.6–6.3] | 5.6 [5.0–6.2] | 6.2 [5.7–6.6] |
| Main type of cigarettes smoked<sup>1</sup> |  |  |  |
| Manufactured | 49.7 [48.9–50.4] | 51.6 [50.4–52.9] | 48.5 [47.5–49.4] |
| Hand-rolled | 50.3 [49.6–51.1] | 48.4 [47.1–49.6] | 51.5 [50.6–52.5] |
| Strength of urges to smoke |  |  |  |
| Mean (SD) | 1.7 (1.3) | 1.6 (1.3) | 1.7 (1.2) |
| 0 Not at all | 23.2 [22.6–23.8] | 28.1 [27.0–29.2] | 20.3 [19.6–21.0] |
| 1 Slight | 20.5 [19.9–21.1] | 18.7 [17.8–19.7] | 21.6 [20.8–22.3] |
| 2 Moderate | 34.8 [34.1–35.5] | 31.7 [30.6–32.9] | 36.6 [35.8–37.5] |
| 3 Strong | 13.7 [13.2–14.2] | 13.7 [12.9–14.6] | 13.7 [13.0–14.3] |
| 4 Very strong | 4.7 [4.4–5.0] | 5.0 [4.5–5.5] | 4.5 [4.1–4.9] |
| 5 Extremely strong | 3.1 [2.9–3.4] | 2.8 [2.4–3.2] | 3.4 [3.1–3.7] |
| Motivation to stop smoking<sup>2,3</sup> |  |  |  |
| Mean (SD) | 3.2 (2.0) | 4.4 (1.8) | 2.7 (1.8) |
| 1 (lowest) | 27.2 [26.5–27.8] | 6.9 [6.2–7.6] | 35.6 [34.7–36.5] |
| 2 | 19.0 [18.4–19.6] | 12.4 [11.4–13.4] | 21.7 [21.0–22.5] |
| 3 | 12.8 [12.2–13.3] | 11.8 [10.8–12.8] | 13.2 [12.6–13.8] |
| 4 | 9.8 [9.4–10.3] | 15.9 [14.9–17.0] | 7.3 [6.9–7.8] |
| 5 | 16.0 [15.4–16.6] | 20.3 [19.1–21.5] | 14.2 [13.6–14.8] |
| 6 | 8.7 [8.2–9.1] | 17.1 [16.0–18.2] | 5.2 [4.8–5.6] |
| 7 (highest) | 6.6 [6.2–7.0] | 15.8 [14.7–16.8] | 2.8 [2.5–3.1] |

<sup>1</sup> Among those who smoked cigarettes in the past year; excludes those who exclusively smoked non-cigarette tobacco.
<sup>2</sup> Among those who reported current smoking; excludes those who successfully quit in the past year.
<sup>3</sup> Response options: 1 = I don't want to stop smoking; 2 = I think I should stop smoking but don't really want to; 3 = I want to stop smoking but haven't thought about when; 4 = I really want to stop smoking but I don't know when I will; 5 = I want to stop smoking and hope to soon; 6 = I really want to stop smoking and intend to in the next 3 months; 7 = I really want to stop smoking and intend to in the next month.

**Table 3.** Prevalence and odds of past-year smoking quit attempts, overall and by sociodemographic characteristics, mental health, and alcohol use.

|  | Tried to quit smoking in the past year |  |  |
| --- | --- | --- | --- |
|  | Row % [95% CI] | OR [95% CI]<sup>1</sup> | OR [95% CI]<sup>2</sup> |
| Age (years) |  |  |  |
| 18-24 | 48.0 [46.2–49.9] | Ref | Ref |
| 25-34 | 41.8 [40.3–43.4] | 0.78 [0.70–0.85] | 0.77 [0.70–0.85] |
| 35-44 | 35.6 [33.9–37.2] | 0.60 [0.54–0.66] | 0.59 [0.53–0.66] |
| 45-54 | 31.1 [29.4–32.7] | 0.49 [0.44–0.54] | 0.48 [0.43–0.54] |
| 55-64 | 31.5 [29.9–33.2] | 0.50 [0.45–0.55] | 0.49 [0.44–0.55] |
| ≥65 | 24.6 [23.1–26.1] | 0.35 [0.31–0.39] | 0.35 [0.31–0.39] |
| Gender |  |  |  |
| Man | 36.0 [35.1–37.0] | Ref | Ref |
| Woman | 37.5 [36.5–38.6] | 1.07 [1.00–1.13] | 1.10 [1.04–1.17] |
| Other | 35.4 [30.5–40.6] | 0.97 [0.78–1.22] | 0.80 [0.63–1.01] |
| Ethnicity |  |  |  |
| White | 36.0 [35.3–36.8] | Ref | Ref |
| Black | 42.4 [38.4–46.4] | 1.31 [1.11–1.55] | 1.15 [0.97–1.37] |
| Asian | 44.0 [40.6–47.5] | 1.40 [1.21–1.62] | 1.24 [1.07–1.44] |
| Mixed/multiple | 39.4 [35.9–43.0] | 1.15 [0.99–1.34] | 0.96 [0.82–1.12] |
| Other | 37.9 [32.7–43.3] | 1.08 [0.86–1.36] | 0.96 [0.76–1.22] |
| Social grade |  |  |  |
| AB (most advantaged) | 35.3 [33.7–36.9] | Ref | Ref |
| C1 | 37.1 [36.1–38.2] | 1.08 [0.99–1.17] | 1.03 [0.94–1.12] |
| C2 | 36.7 [35.2–38.2] | 1.06 [0.96–1.17] | 1.04 [0.95–1.15] |
| D | 38.6 [36.5–40.7] | 1.15 [1.02–1.28] | 1.10 [0.98–1.23] |
| E (least advantaged) | 35.2 [33.6–36.9] | 1.00 [0.90–1.10] | 1.11 [1.01–1.24] |
| Out of work due to long-term ill health or disability |  |  |  |
| No | 36.6 [35.9–37.3] | Ref | Ref |
| Yes | 37.8 [35.6–40.2] | 1.06 [0.96–1.17] | 1.14 [1.01–1.28] |
| Diagnosed mental health condition |  |  |  |
| No | 34.7 [33.5–36.0] | Ref | Ref |
| Yes | 40.3 [38.9–41.7] | 1.27 [1.17–1.38] | 1.19 [1.09–1.30] |
| Past-month psychological distress |  |  |  |
| None/low | 33.9 [32.7–35.1] | Ref | Ref |
| Moderate | 40.1 [38.5–41.7] | 1.31 [1.20–1.43] | 1.14 [1.04–1.24] |
| Severe | 41.6 [39.2–43.9] | 1.39 [1.25–1.56] | 1.17 [1.03–1.32] |
| Alcohol consumption |  |  |  |
| None | 38.0 [36.6–39.4] | Ref | Ref |
| Low risk | 38.1 [36.8–39.4] | 1.00 [0.92–1.09] | 0.99 [0.91–1.07] |
| Increasing/higher risk | 35.1 [34.0–36.2] | 0.88 [0.82–0.95] | 0.80 [0.74–0.87] |

CI, confidence interval. OR, odds ratio.<sup>1</sup> Odds of having made a past-year smoking quit attempt, adjusted for survey year.
<sup>2</sup> Odds of having made a past-year smoking quit attempt, adjusted for survey year, age, gender, ethnicity, and social grade.

**Table 4.**
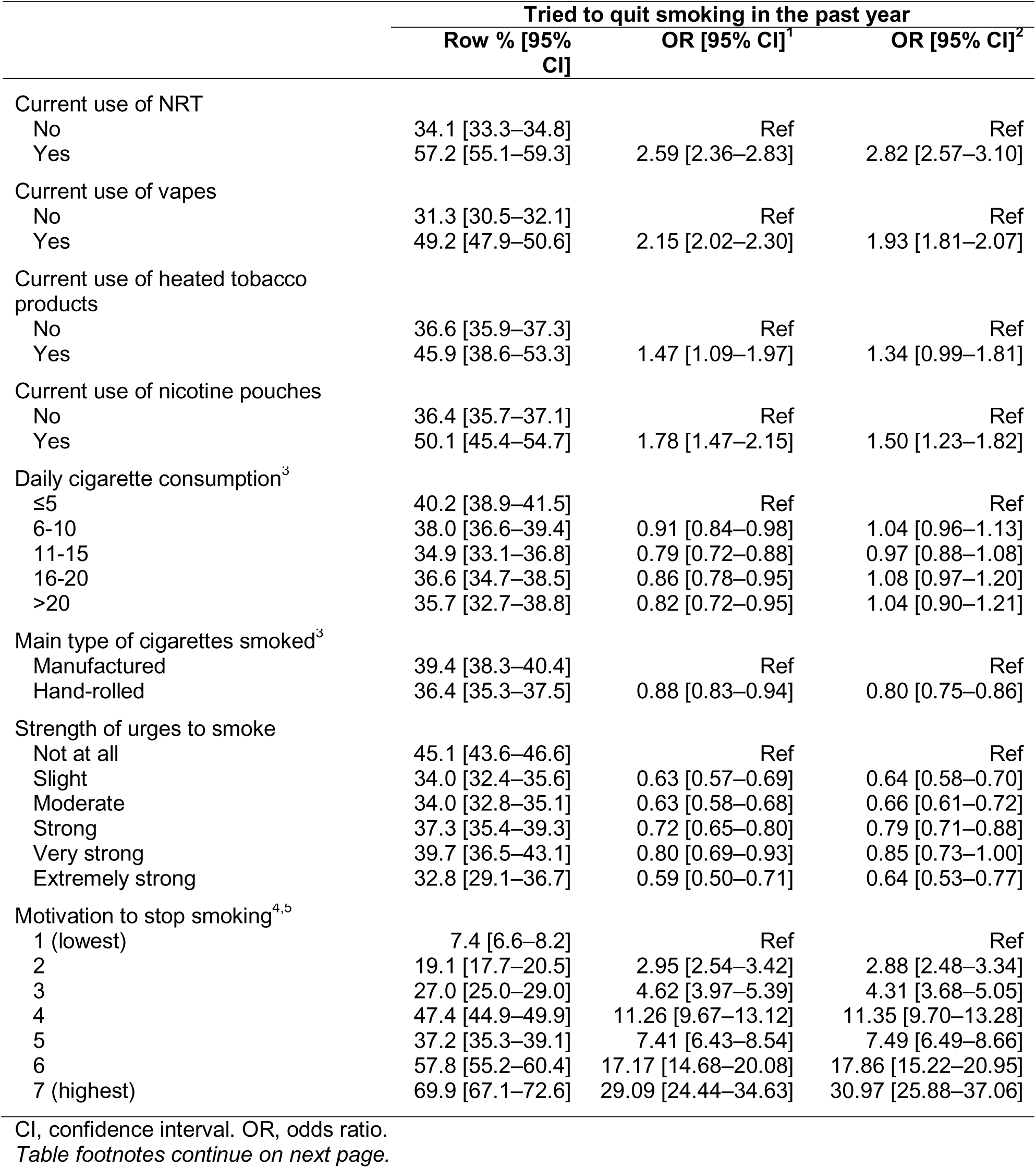
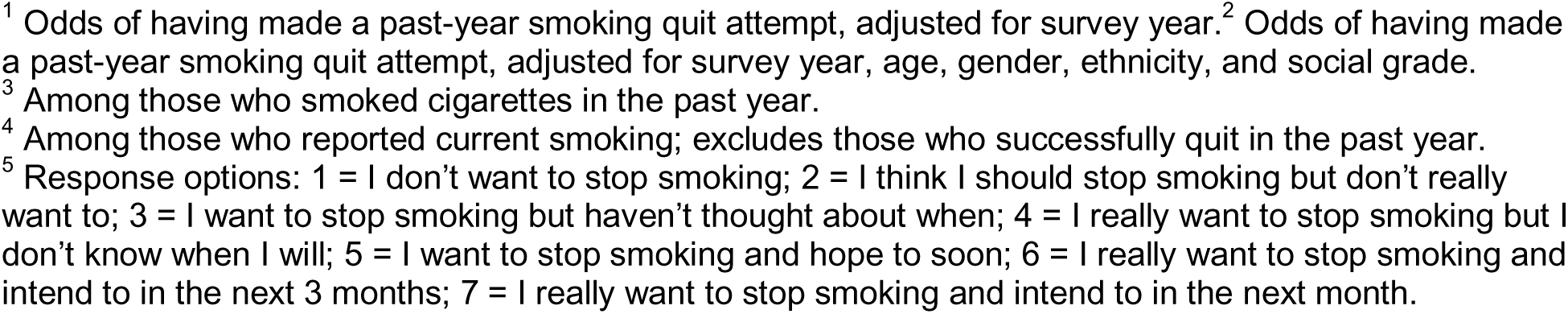
Prevalence and odds of past-year smoking quit attempts, by non-combustible nicotine use and smoking characteristics.

|  | Tried to quit smoking in the past year |  |  |
| --- | --- | --- | --- |
|  | Row % [95% CI] | OR [95% CI]<sup>1</sup> | OR [95% CI]<sup>2</sup> |
| Current use of NRT |  |  |  |
| No | 34.1 [33.3–34.8] | Ref | Ref |
| Yes | 57.2 [55.1–59.3] | 2.59 [2.36–2.83] | 2.82 [2.57–3.10] |
| Current use of vapes |  |  |  |
| No | 31.3 [30.5–32.1] | Ref | Ref |
| Yes | 49.2 [47.9–50.6] | 2.15 [2.02–2.30] | 1.93 [1.81–2.07] |
| Current use of heated tobacco products |  |  |  |
| No | 36.6 [35.9–37.3] | Ref | Ref |
| Yes | 45.9 [38.6–53.3] | 1.47 [1.09–1.97] | 1.34 [0.99–1.81] |
| Current use of nicotine pouches |  |  |  |
| No | 36.4 [35.7–37.1] | Ref | Ref |
| Yes | 50.1 [45.4–54.7] | 1.78 [1.47–2.15] | 1.50 [1.23–1.82] |
| Daily cigarette consumption<sup>3</sup> |  |  |  |
| ≤5 | 40.2 [38.9–41.5] | Ref | Ref |
| 6-10 | 38.0 [36.6–39.4] | 0.91 [0.84–0.98] | 1.04 [0.96–1.13] |
| 11-15 | 34.9 [33.1–36.8] | 0.79 [0.72–0.88] | 0.97 [0.88–1.08] |
| 16-20 | 36.6 [34.7–38.5] | 0.86 [0.78–0.95] | 1.08 [0.97–1.20] |
| >20 | 35.7 [32.7–38.8] | 0.82 [0.72–0.95] | 1.04 [0.90–1.21] |
| Main type of cigarettes smoked<sup>3</sup> |  |  |  |
| Manufactured | 39.4 [38.3–40.4] | Ref | Ref |
| Hand-rolled | 36.4 [35.3–37.5] | 0.88 [0.83–0.94] | 0.80 [0.75–0.86] |
| Strength of urges to smoke |  |  |  |
| Not at all | 45.1 [43.6–46.6] | Ref | Ref |
| Slight | 34.0 [32.4–35.6] | 0.63 [0.57–0.69] | 0.64 [0.58–0.70] |
| Moderate | 34.0 [32.8–35.1] | 0.63 [0.58–0.68] | 0.66 [0.61–0.72] |
| Strong | 37.3 [35.4–39.3] | 0.72 [0.65–0.80] | 0.79 [0.71–0.88] |
| Very strong | 39.7 [36.5–43.1] | 0.80 [0.69–0.93] | 0.85 [0.73–1.00] |
| Extremely strong | 32.8 [29.1–36.7] | 0.59 [0.50–0.71] | 0.64 [0.53–0.77] |
| Motivation to stop smoking<sup>4,5</sup> |  |  |  |
| 1 (lowest) | 7.4 [6.6–8.2] | Ref | Ref |
| 2 | 19.1 [17.7–20.5] | 2.95 [2.54–3.42] | 2.88 [2.48–3.34] |
| 3 | 27.0 [25.0–29.0] | 4.62 [3.97–5.39] | 4.31 [3.68–5.05] |
| 4 | 47.4 [44.9–49.9] | 11.26 [9.67–13.12] | 11.35 [9.70–13.28] |
| 5 | 37.2 [35.3–39.1] | 7.41 [6.43–8.54] | 7.49 [6.49–8.66] |
| 6 | 57.8 [55.2–60.4] | 17.17 [14.68–20.08] | 17.86 [15.22–20.95] |
| 7 (highest) | 69.9 [67.1–72.6] | 29.09 [24.44–34.63] | 30.97 [25.88–37.06] |

CI, confidence interval. OR, odds ratio.
Table footnotes continue on next page.
<sup>1</sup> Odds of having made a past-year smoking quit attempt, adjusted for survey year.<sup>2</sup> Odds of having made a past-year smoking quit attempt, adjusted for survey year, age, gender, ethnicity, and social grade.
<sup>3</sup> Among those who smoked cigarettes in the past year.
<sup>4</sup> Among those who reported current smoking; excludes those who successfully quit in the past year.
<sup>5</sup> Response options: 1 = I don't want to stop smoking; 2 = I think I should stop smoking but don't really want to; 3 = I want to stop smoking but haven't thought about when; 4 = I really want to stop smoking but I don't know when I will; 5 = I want to stop smoking and hope to soon; 6 = I really want to stop smoking and intend to in the next 3 months; 7 = I really want to stop smoking and intend to in the next month.

### Overall and annual prevalence of quit attempts

Overall, 36.7% [95% CI 36.0–37.4] of participants reported making at least one smoking quit attempt in the past year, while 63.3% [62.6–64.0] reported no attempts. The annual prevalence of quit attempts was relatively stable over the study period (range: 35.5% [33.9–37.1] to 37.6% [35.9–39.3]), as was the estimated number of adults attempting to quit smoking each year (range: 3.45 [3.15–3.76] to 3.73 [3.56–3.89] million; **Table S2**).

### Associations with sociodemographic and socioeconomic variables

Quit attempts were most common among younger adults and declined progressively with age, from 48.0% among those aged 18–24 years to 24.6% among those aged ≥65 years (**Table 3**). Relative to those aged 18–24 years, all older age groups had lower odds of reporting a quit attempt, with the lowest odds observed among adults aged ≥65 years (OR=0.35, 95% CI 0.31– 0.39). Half (50.3%) of participants who reported making a quit attempt were aged under 35 years (**Table 1**).

There was little evidence of substantial variation in quit attempts by gender, social grade, or health-related economic inactivity (**Table 3**). Those reporting health-related economic inactivity had slightly higher odds of making a quit attempt after adjustment for age, gender, ethnicity, and social grade (OR=1.14, 95% CI 1.01–1.28), but the 95% CI included the possibility of no difference (**Table 3**).

Differences were observed by ethnicity (**Table 3**), with quit attempts more common among Black (42.4%) and Asian adults (44.0%) than White adults (36.0%; OR=1.31, 95% CI 1.11–1.55 and OR=1.40, 95% CI 1.21–1.62, respectively). These patterns were partly explained by other sociodemographic variables, attenuating to OR=1.15 (95% CI 0.97–1.37) and OR=1.24 (95% CI 1.07–1.44), respectively after adjustment for age, gender, and social grade (**Table 3**).

### Associations with mental health indicators and alcohol use

Adults with indicators of poorer mental health were more likely to report a quit attempt (**Table 3**). The prevalence of quit attempts was higher among those with vs. without a diagnosed mental health condition (40.3% vs. 34.7%; OR=1.27, 95% CI 1.17–1.38) and higher with greater levels of past-month psychological distress, ranging from 33.9% among those reporting no/low distress to 41.6% among those reporting severe distress (OR=1.39, 95% CI 1.25–1.56). Half (49.8%) of participants who reported making a quit attempt reported a diagnosed mental health condition and just over half (52.8%) reported experiencing moderate or severe psychological distress, compared with 43.9% and 45.6%, respectively, among those who did not attempt to quit (**Table 1**).

These associations were partly explained by sociodemographic and socioeconomic differences between groups. After adjustment for age, gender, ethnicity, and social grade, associations attenuated to OR=1.19 (95% CI 1.09–1.30) for those with vs. without diagnosed mental health conditions, and to OR=1.14 (95% CI 1.04–1.24) and OR=1.17 (95% CI 1.03–1.32) for those with moderate and severe distress vs. no/low distress (**Table 3**).

There were also differences by alcohol consumption (**Table 3**), with adults reporting increasing/higher-risk consumption being slightly less likely to report a quit attempt than non-drinkers (35.1% vs. 38.0%; OR=0.88, 95% CI 0.82–0.95). This pattern persisted after adjustment for sociodemographic and socioeconomic characteristics (**Table 3**).

### Associations with non-combustible nicotine use and smoking-related characteristics

Quit attempts also differed according to use of non-combustible nicotine products and smoking-related characteristics (**Table 4**).

Current use of non-combustible nicotine products at the time of the survey was associated with higher prevalence of quit attempts, particularly among people currently using NRT (57.2%; OR=2.59, 95% CI 2.36–2.83) and vapes (49.2%; OR=2.15, 95% CI 2.02–2.30; **Table 4**). These patterns held after adjustment for sociodemographic variables (**Table 4**) and when the sample was restricted to those reporting current smoking (**Table S5**). Among those who had made a past-year quit attempt, 17.6% reported current use of NRT, 40.4% vapes, 3.3% nicotine pouches, and 1.3% heated tobacco products, compared with 7.6%, 24.2%, 1.9%, and 0.9% among those who did not attempt to quit (**Table 2**).

Associations with indicators of dependence varied across models (**Table 4; Table S5**). Among all adults who had smoked in the past year, those who reported smoking >5 cigarettes per day had lower odds of making a past-year quit attempt than those who reported smoking ≤5 cigarettes per day (OR range 0.79–0.91), but this association between daily cigarette consumption and quit attempts was attenuated after adjustment for sociodemographic factors (OR range 0.97–1.08; **Table 4**). This largely reflected differences in age across consumption groups: those reporting lower consumption were younger on average than those who smoked more heavily (mean age = 37.0 years among those smoking ≤5 cigarettes/day, 43.3 years among those smoking 5-10, and 46.5-47.2 years among those reporting higher consumption). When we restricted the sample to adults who currently smoked, there was a clearer gradient whereby making a quit attempt was less common at higher levels of consumption (OR range 0.59–0.86); a pattern that was attenuated but not completely explained by adjustment for sociodemographic variables (OR range 0.69–0.95; **Table S5**).

Among all adults who had smoked in the past year, reporting stronger urges to smoke in the past 24 hours was associated with reduced odds of having made a past-year quit attempt, with the lowest odds observed among those reporting extremely strong urges (OR=0.59, 95% CI 0.50–0.71; **Table 4**). However, results differed when we restricted the analysis to adults who currently smoked, among whom making a past-year quit attempt was least common in those reporting no urges to smoke in the past 24 hours, with higher odds among those reporting any urges (OR range 1.68–2.55) and highest among those reporting very strong urges (OR=2.55, 95% CI 2.14–3.03; **Table S5**). For both denominators, these patterns persisted after adjustment for sociodemographic factors (**Table 4; Table S5**).

Adults who mainly smoked hand-rolled cigarettes were also slightly less likely to report a quit attempt than those who smoked manufactured cigarettes (36.4% vs. 39.4%; OR=0.88, 95% CI 0.83–0.94; **Table 4**). This pattern was observed consistently when we adjusted for sociodemographic variables (**Table 4**) and restricted the sample to those reporting current smoking (**Table S5**).

Among adults who reported current smoking, there were stark differences in the prevalence of past-year quit attempts by motivation to stop smoking (**Table 4**), ranging from 7.4% among those who said they did not want to stop smoking to 69.9% among those who said they really wanted to stop smoking and intended to in the next month (OR=29.09, 95% CI 24.44–34.63). This pattern persisted after adjustment for sociodemographic variables (**Table 4**). Among those who currently smoked and had made a past-year quit attempt, one in three (32.9%) said they really wanted to stop smoking and intended to in the next 1-3 months, compared with 8.0% of those who had not made a quit attempt (**Table 2**).

### Geographic variation

Geographic differences were modest (**Figure 1; Table S3**). The national prevalence of quit attempts was highest in England (37.0%) and lowest in Wales (32.1%; OR=0.80, 95% CI 0.72– 0.89), with Scotland in between (35.4%). Within England, regional prevalence of quit attempts ranged from 35.4% in the East of England to 39.4% in the North East, with CIs overlapping for all regions (**Table S3**).

**Figure 1.**
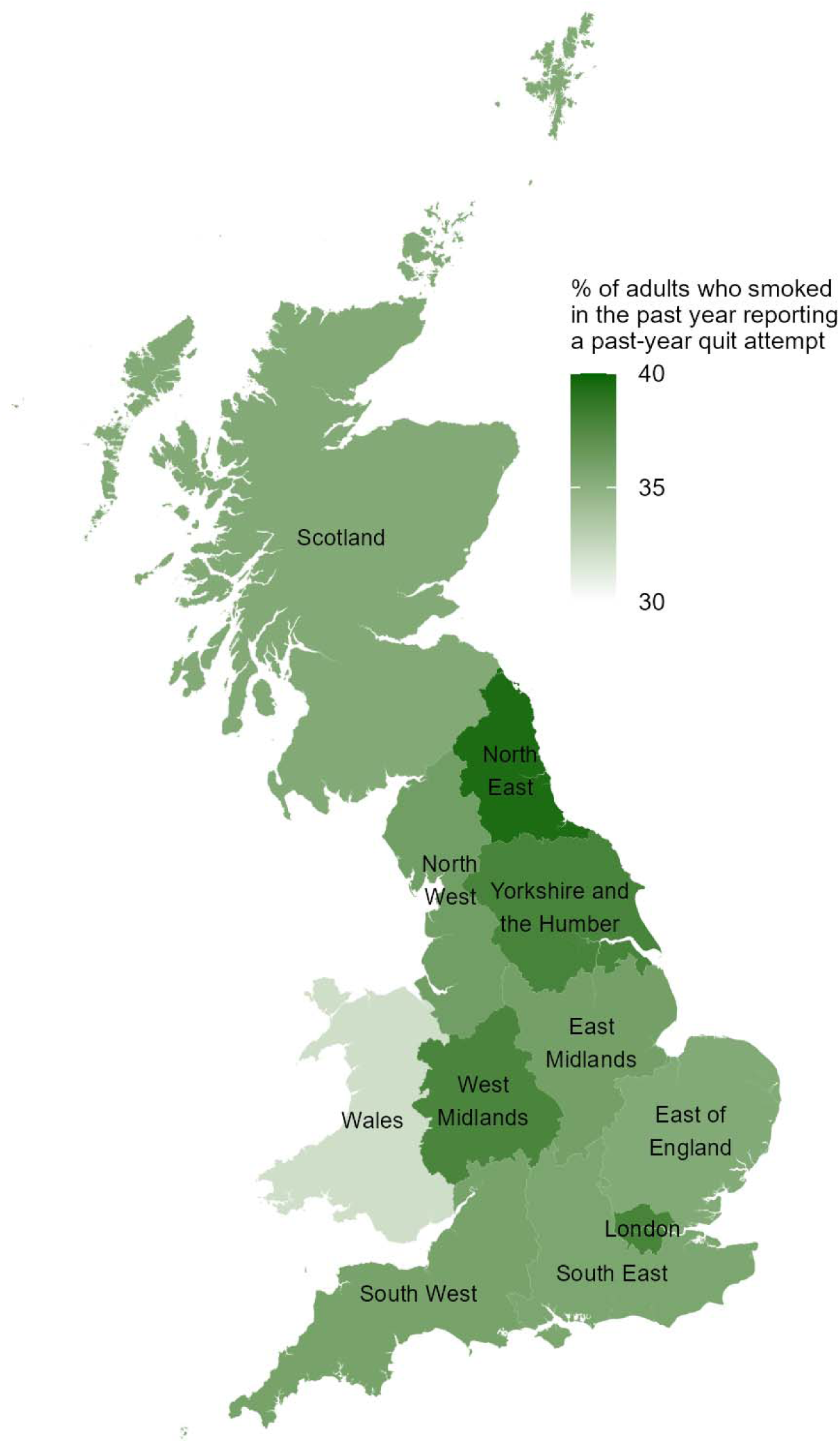
Prevalence of past-year smoking quit attempts in Scotland, Wales, and regions of England. Prevalence estimates and odds ratios are reported in **Table S3**.

### Interactions with survey year

Tests of interactions with survey year provided no clear evidence that associations with participant characteristics or geographic area varied over time (**Table S4**).

## Discussion

In this nationally representative study of adults in Great Britain who had smoked tobacco in the past year, just over one in three reported making at least one serious quit attempt in the past-year, corresponding to approximately 3.5 million people per year. This estimate was broadly stable between 2020 and 2026, suggesting no clear change in quit attempt prevalence during the post-pandemic period despite ongoing developments in tobacco control policy, cost-of-living pressures, and expanded cessation support provision.

The estimated absolute number of adults making smoking quit attempts may appear high relative to commonly cited estimates of the smoking population (approximately 5–6 million adults in Great Britain).^3^ However, this reflects differences in denominators: our estimate includes all adults who had smoked tobacco in the past year (including those who successfully quit within the past 12 months), whereas national prevalence estimates (e.g., Annual Population Survey) are restricted to current cigarette smoking, may underrepresent people who smoke non-daily, and do not capture those who exclusively smoke non-cigarette tobacco.^41,42^

Consistent with previous work,^16,19,20^ our data showed greater variation in quit attempts by smoking-related characteristics than by sociodemographic factors. Motivation to stop smoking showed a strong positive gradient among those who currently smoked. This aligns with established models of behaviour change, in which motivation is a strong predictor of taking action.^43^ Associations with indicators of nicotine dependence were more nuanced. The complete attenuation of the association between cigarette consumption and quit attempts after adjustment in the full sample, but not among adults who currently smoked, suggests that this relationship is influenced both by differences in the sociodemographic profile of people with different levels of cigarette consumption and by who is successful in quitting. By contrast, the reversal in the association with current urges to smoke highlights the importance of temporality: current urges among people who had already quit are likely to have been reduced by abstinence and therefore may not reflect urges at the time of the quit attempt. Together, these findings suggest that measures of nicotine dependence should be interpreted cautiously when examining past quit attempts, particularly in samples that include both adults who currently smoke and those who have successfully quit.

Use of hand-rolled cigarettes was associated with slightly lower odds of quitting. This may reflect differences in smoking patterns and dependence, including the ritualised aspects of preparing and smoking hand-rolled cigarettes,^44,45^ which may reinforce behavioural attachment to smoking. Lower cost of hand-rolled cigarettes^46^ may also reduce financial incentives to quit for some people, although cost has been identified as an increasingly important motive for smoking cessation in recent years.^47^ Use of non-combustible nicotine products, particularly NRT and vapes, was strongly associated with reporting a quit attempt. This likely reflects reverse causality, whereby individuals who initiate quit attempts are more likely to use cessation aids rather than these products independently increasing quit attempt rates.

Although weaker than associations with smoking-related variables, there were some notable sociodemographic patterns. Age showed a clear gradient, with progressively lower rates of quit attempts in older age groups. This is consistent with some prior evidence^18^ and may reflect differences in perceived benefits of quitting, accumulated dependence, responsiveness to health-related prompts, or self-selection whereby those who are still smoking at older ages are more likely to be more heavily dependent^48^ or less motivated to quit.^49^ This may in turn be related to lower perceived urgency or perceived benefit of quitting among older adults who have smoked for longer periods without experiencing major smoking-related health consequences.

This highlights the importance of ensuring that tobacco control efforts do not exclusively focus on young people, but that cessation campaigns and support services effectively engage older adults, who may be less likely to make quit attempts despite substantial potential health gains from cessation.^50^ We also observed higher rates of quit attempts among Black and Asian adults than among White adults, although the reasons for these differences are unclear and warrant further investigation. In contrast, associations with gender, social grade, and health-related economic inactivity were small or inconsistent.

Importantly, individuals with diagnosed mental health conditions and those reporting higher psychological distress were more likely to report a quit attempt. Associations were attenuated but remained after adjustment for sociodemographic characteristics, suggesting that differences in age, gender, ethnicity, and social grade account for only part of the higher likelihood of quit attempts among those with poorer mental health. This pattern is consistent with previous research showing high levels of motivation to stop smoking among people with mental health conditions,^51^ despite their higher rates of smoking and greater dependence.^52–55^ Several explanations are plausible. Greater symptom burden^56,57^ and more frequent contact with health and social care services may increase exposure to cessation advice and strengthen motivation to stop smoking, even in the presence of higher dependence. Distress may also make health concerns more of a priority. However, these findings should be interpreted cautiously, as psychological distress may also arise following unsuccessful quit attempts, relapse, or nicotine withdrawal. Together with the lack of a clear gradient by social grade, these findings are consistent with the interpretation that inequalities in smoking prevalence are driven less by differences in quit attempt initiation and more by differences in smoking uptake, quit success, or relapse.^58^

Alcohol use showed a modest inverse association with quit attempts, although the magnitude of association was small. This differs from some previous research, which found no evidence that drinkers at risk of alcohol dependence differed in their likelihood of making a past-year quit attempt compared with lower-risk drinkers, although higher-risk drinking was associated with lower quit success.^59^ Geographic variation in quit attempts was modest, suggesting relatively limited regional differences in quit initiation behaviour within Great Britain compared with stronger individual-level variation. We note that the rate of quit attempts was highest in the North East of England, which could be linked to its strong regional tobacco control programme,^60^ although this cross-sectional analysis cannot establish causal effects. It is also possible that broad regional analyses mask important local variation, as deprived and more affluent neighbourhoods may coexist within the same geographic area. Future research should therefore examine smaller-area variation to identify communities with particularly low quit attempt rates and unmet cessation needs.

Strengths of this study include the large, nationally representative sample and detailed assessment of sociodemographic, socioeconomic, and smoking-related variables. Several limitations should be noted. First, all measures were self-reported and therefore subject to recall and reporting bias, particularly for variables that captured past behaviour among participants who had successfully quit smoking in the past year. Second, some measures were not necessarily reflective of participants’ circumstances at the time of their quit attempt. Among those who had quit within the past year, cigarette consumption reflected previous smoking behaviour, while current non-combustible nicotine use and levels of psychological distress may have changed following the quit attempt. Motivation to stop smoking was assessed only among current smokers, limiting comparisons with those who had successfully quit. Third, the cross-sectional design precludes causal inference, and associations (e.g., between non-combustible nicotine use and quit attempts) likely reflect bidirectional relationships and confounding by underlying motivation to quit. Fourth, as a household-based survey, the sample underrepresents certain populations, including individuals not living in private households (e.g. institutionalised populations or those experiencing homelessness).^61^ Finally, although we examined a range of sociodemographic, mental health, and smoking-related characteristics separately, we did not assess their intersection. As a result, we were unable to identify whether certain combinations of characteristics (e.g., older age combined with high dependence and socioeconomic disadvantage) define subgroups with particularly low likelihood of attempting to quit. Future research using intersectional approaches (e.g., MAIHDA) could help identify strata with particularly low rates of quit attempts, which may be important for more targeted cessation interventions and public health messaging.

In conclusion, approximately one-third of adults who smoked in the past year reported a quit attempt in the post-pandemic period, with stable prevalence over this period. Variation in quit attempts was driven more by motivation to quit and nicotine dependence than by sociodemographic or socioeconomic characteristics. Individuals least likely to attempt quitting tended to be characterised by lower motivation to quit, greater nicotine dependence, and older age. These findings support the interpretation that inequalities in smoking prevalence are unlikely to be sustained by differences in the initiation of quit attempts, but rather by differences in success and relapse once attempts are made. Efforts to reduce smoking prevalence should focus on increasing quit attempts among groups less likely to try, while recognising that reducing inequalities will also require attention to access effective support, increase quit success, and reduce relapse back to smoking.

## Data Availability

All data produced in the present study are available upon reasonable request to the authors.

## Declarations

### Ethics approval

Ethical approval for the STS was granted originally by the UCL Ethics Committee (ID 0498/001). Participants provide informed consent to take part in the study, and all methods are carried out in accordance with relevant regulations. The data are not collected by UCL and are anonymised when received by UCL.

