## Supplementary file for "Who is and who is not attempting to quit smoking? A population study in Great Britain, 2020–2026"

**Table S1.** Sample characteristics

| | Unweighted *n* | Valid column % |
|---|---|---|
| Age (years) | | |
| 18-24 | 3,630 | 16.9 |
| 25-34 | 5,253 | 24.7 |
| 35-44 | 4,022 | 18.0 |
| 45-54 | 3,937 | 15.5 |
| 55-64 | 3,840 | 12.9 |
| ≥65 | 4,104 | 12.0 |
| *Missing* | 0 | - |
| Gender | | |
| Man | 13,124 | 53.2 |
| Woman | 11,212 | 45.6 |
| Other | 342 | 1.2 |
| *Missing* | 108 | - |
| Ethnicity | | |
| White | 21,500 | 87.2 |
| Black | 729 | 3.0 |
| Asian | 1,023 | 4.3 |
| Mixed/multiple | 928 | 3.9 |
| Other | 402 | 1.6 |
| *Missing* | 204 | - |
| Social grade | | |
| AB (most advantaged) | 4,173 | 16.3 |
| C1 | 9,452 | 26.1 |
| C2 | 4,612 | 25.0 |
| D | 2,571 | 17.5 |
| E (least advantaged) | 3,978 | 15.1 |
| *Missing* | 0 | - |
| Out of work due to long-term ill health or disability | | |
| No | 22,528 | 91.1 |
| Yes | 2,173 | 8.9 |
| *Missing* | 85 | - |
| Nation | | |
| England | 18,518 | 88.4 |
| Wales | 2,155 | 4.1 |
| Scotland | 4,113 | 7.5 |
| *Missing* | 0 | - |
| Diagnosed mental health condition[1] | | |
| No | 6,780 | 53.9 |
| Yes | 5,599 | 46.1 |
| *Missing*[1] | 1,903 | - |

*Table continues on next page.*

**Table S1.** *Continued*

| | **Unweighted *n*** | **Valid column %** |
|---|---|---|
| Past-month psychological distress[1] | | |
| None/low | 7,324 | 51.7 |
| Moderate | 4,345 | 32.7 |
| Severe | 2,024 | 15.6 |
| *Missing*[1] | 589 | - |
| Alcohol consumption | | |
| None | 6,391 | 27.9 |
| Low risk | 6,913 | 30.0 |
| Increasing/higher risk | 9,714 | 42.1 |
| *Missing* | 1,768 | - |
| Current use of… | | |
| Nicotine replacement therapy | 2,902 | 11.3 |
| Vapes | 7,005 | 30.1 |
| Heated tobacco products | 236 | 1.0 |
| Nicotine pouches | 561 | 2.5 |
| *Missing* | 0 | - |
| Daily cigarette consumption[2] | | |
| ≤5 | 7,338 | 35.0 |
| 6-10 | 6,106 | 28.9 |
| 11-15 | 3,454 | 15.3 |
| 16-20 | 3,314 | 14.8 |
| >20 | 1,321 | 5.9 |
| *Missing*[2] | 1,166 | - |
| Main type of cigarettes smoked[2] | | |
| Manufactured | 11,104 | 49.7 |
| Hand-rolled | 9,889 | 50.3 |
| *Missing*[2] | 1,706 | - |
| Strength of urges to smoke | | |
| Not at all | 5,509 | 23.2 |
| Slight | 4,815 | 20.5 |
| Moderate | 8,406 | 34.8 |
| Strong | 3,303 | 13.7 |
| Very strong | 1,109 | 4.7 |
| Extremely strong | 773 | 3.1 |
| *Missing* | 871 | - |

[1] Assessed in a subset of waves; see measures. Numbers of missing cases exclude participants surveyed in other waves (*n*=8,860) and participants in Wales (*n*=553) and Scotland (*n*=1,091) who were not asked these questions.
[2] Among those who smoked cigarettes in the past year. The number of missing cases excludes those who currently exclusively smoked non-cigarette tobacco (*n*=2,087) but includes an unknown number who exclusively smoked non-cigarette tobacco but successfully quit in the past year.

**Table S2.** Annual prevalence of past-year smoking quit attempts among adults in Great Britain who reported smoking in the past year

| | Year[1] | | | | | | |
|---|---|---|---|---|---|---|---|
| | **2020** | **2021** | **2022** | **2023** | **2024** | **2025** | **2026** |
| Total population size, *n* (millions; ONS)[2*] | 51.42 | 51.69 | 52.20 | 52.91 | 53.53 | 53.53* | 53.53* |
| Past-year smoking prevalence, % [95% CI] (STS) | 18.2 [17.2–19.3] | 18.8 [18.3–19.4] | 19.3 [18.8–19.9] | 18.8 [18.2–19.3] | 18.4 [17.8–19.0] | 19.2 [18.6–19.7] | 17.6 [16.8–18.5] |
| Past-year smoking quit attempt, % [95% CI] (STS) | | | | | | | |
| Yes | 36.9 [33.7–40.2] | 37.4 [35.8–39.0] | 37.0 [35.3–38.6] | 35.9 [34.3–37.6] | 37.6 [35.9–39.3] | 35.5 [33.9–37.1] | 37.0 [34.3–39.7] |
| No | 63.1 [59.8–66.3] | 62.6 [61.0–64.2] | 63.0 [61.4–64.7] | 64.1 [62.4–65.7] | 62.4 [60.7–64.1] | 64.5 [62.9–66.1] | 63.0 [60.3–65.7] |
| Number of adults who made a past-year smoking quit attempt, *n* (millions)[3] | 3.45 [3.15–3.76] | 3.63 [3.48–3.79] | 3.73 [3.56–3.89] | 3.57 [3.41–3.74] | 3.70 [3.54–3.87] | 3.65* [3.48–3.81] | 3.49* [3.23–3.74] |

ONS, Office for National Statistics. STS, Smoking Toolkit Study.

[1] Note: 2020 includes data from October-December only and 2026 from January-May only.

[2] Mid-year population estimate: number of adults aged ≥18 years in Great Britain.

* In the absence of published mid-year population size estimates for 2025 and 2026, these numbers use the mid-year estimate for 2024 and therefore likely slightly underestimate actual numbers for 2025 and 2026.

[3] Calculated as the total number of adults in Great Britain x proportion who smoked in the past year x proportion of those who smoked in the past year who made a smoking quit attempt. Confidence intervals were derived by repeating the calculation using the lower and upper 95% confidence limits for the proportion of those who smoked in the past year who made a smoking quit attempt.

**Table S3.** Prevalence and odds of past-year smoking attempts, by geographic area

| | **Tried to quit smoking in the past year** | |
|---|---|---|
| | **Row % [95% CI]** | **OR [95% CI][1]** |
| Nation | | |
| England | 37.0 [36.2–37.8] | Ref |
| Wales | 32.1 [29.9–34.3] | 0.80 [0.72–0.89] |
| Scotland | 35.4 [33.8–37.0] | 0.93 [0.72–0.89] |
| Region of England | | |
| North East | 39.4 [35.9–43.1] | Ref |
| North West | 36.2 [34.1–38.3] | 0.87 [0.73–1.04] |
| Yorkshire and the Humber | 37.9 [35.5–40.4] | 0.94 [0.78–1.13] |
| East Midlands | 36.0 [33.5–38.7] | 0.87 [0.72–1.05] |
| West Midlands | 37.8 [35.4–40.3] | 0.94 [0.78–1.13] |
| East of England | 35.4 [33.2–37.7] | 0.84 [0.70–1.01] |
| London | 38.0 [36.1–39.8] | 0.94 [0.80–1.12] |
| South East | 35.7 [33.8–37.6] | 0.86 [0.72–1.02] |
| South West | 35.9 [33.6–38.2] | 0.86 [0.72–1.04] |

CI, confidence interval. OR, odds ratio.
[1] Odds of having made a past-year smoking quit attempt, adjusted for survey year.

**Table S4.** Interactions between participant characteristics and survey year

| | *F* | *p* |
|---|---|---|
| Age | 1.18 | 0.224 |
| Gender | 0.68 | 0.775 |
| Ethnicity | 0.99 | 0.470 |
| Social grade | 0.86 | 0.660 |
| Out of work due to long-term ill health or disability | 0.82 | 0.553 |
| Diagnosed mental health condition | 0.69 | 0.602 |
| Past-month psychological distress | 1.20 | 0.288 |
| Alcohol consumption | 0.47 | 0.934 |
| Current use of NRT | 1.83 | 0.090 |
| Current use of vapes | 1.24 | 0.280 |
| Current use of heated tobacco products | 0.92 | 0.479 |
| Current use of nicotine pouches | 0.29 | 0.942 |
| Daily cigarette consumption[1] | 0.79 | 0.749 |
| Main type of cigarettes smoked[1] | 1.19 | 0.309 |
| Strength of urges to smoke | 1.01 | 0.452 |
| Motivation to stop smoking[2] | 0.93 | 0.589 |
| Nation | 1.60 | 0.085 |
| Region in England | 0.83 | 0.787 |

NRT, nicotine replacement therapy.
[1] Among those who smoked cigarettes in the past year.
[2] Among those who reported current smoking; excludes those who successfully quit in the past year.

### Summary of profile comparisons

There were modest differences in sociodemographic characteristics, mental health, and alcohol use between those who did and those who did not report making at least one smoking quit attempt in the past year (**Table 3**). Those who attempted to quit were younger on average (mean age 38.9 vs 44.1 years), with half (50.3%) aged under 35 compared with just over a third (36.6%) among those who did not try to quit. Quit attempts were also slightly less likely to be White (85.6% vs. 88.2%) and more likely to be Asian (5.2% vs. 3.8%). Other sociodemographic characteristics were broadly similar between groups. Quit attempters were slightly more likely to report a diagnosed mental health condition (49.8% vs 43.9%) and moderate (35.3% vs. 31.1%) or severe (17.5% vs. 14.5%) psychological distress. They were also slightly less likely to report increasing- or higher-risk alcohol consumption (40.1% vs. 43.3%).

Marked differences were observed for non-combustible nicotine use (**Table 4**). Those who had attempted to quit were substantially more likely to report current use of nicotine replacement therapy (17.6% vs 7.6%) and vapes (40.4% vs 24.2%), with smaller but consistent differences also observed for heated tobacco products and nicotine pouches.

Differences in cigarette consumption were relatively small (**Table 4**). Mean daily cigarette consumption was similar between groups, although those who attempted to quit were slightly more likely to be lighter smokers (≤5 cigarettes per day: 37.1% vs 33.7%) and slightly less likely to mainly smoke hand-rolled cigarettes (48.4% vs. 51.5%). Quit attempters were more likely to report no urges to smoke in the past 24 hours (28.1% vs. 20.3%) which may reflect some of this group having successfully quit. The proportions reporting strong, very strong, or extremely strong urges were similar across groups.

Among adults who reported current smoking, motivation to stop smoking showed a pronounced gradient (**Table 4**). Those who had made a past-year quit attempt were more likely to report higher levels of motivation than those who had not, particularly at the upper end of the scale (e.g., 32.9% vs. 8.0% said they really wanted to stop smoking and intended to in the next 1-3 months).

**Table S5.** Prevalence and odds of past-year smoking quit attempts, by non-combustible nicotine use and smoking characteristics – among adults who reported current smoking

| | Tried to quit smoking in the past year | | |
|---|---|---|---|
| | **Row % [95% CI]** | **OR [95% CI][1]** | **OR [95% CI][2]** |
| Current use of NRT | | | |
| No | 26.3 [25.5–27.0] | Ref | Ref |
| Yes | 51.8 [49.6–54.1] | 3.02 [2.74–3.34] | 3.30 [2.98–3.65] |
| Current use of vapes | | | |
| No | 24.4 [23.6–25.2] | Ref | Ref |
| Yes | 41.1 [39.6–42.5] | 2.19 [2.04–2.36] | 1.99 [1.84–2.15] |
| Current use of heated tobacco products | | | |
| No | 29.0 [28.3–29.7] | Ref | Ref |
| Yes | 39.5 [32.1–47.5] | 1.60 [1.16–2.21] | 1.47 [1.05–2.05] |
| Current use of nicotine pouches | | | |
| No | 28.8 [28.1–29.5] | Ref | Ref |
| Yes | 43.6 [38.6–48.7] | 1.97 [1.60–2.43] | 1.71 [1.38–2.12] |
| Daily cigarette consumption[3] | | | |
| ≤5 | 33.3 [32.0–34.7] | Ref | Ref |
| 6-10 | 30.1 [28.7–31.5] | 0.86 [0.78–0.94] | 0.95 [0.87–1.05] |
| 11-15 | 28.6 [26.7–30.5] | 0.79 [0.71–0.89] | 0.93 [0.82–1.04] |
| 16-20 | 25.9 [24.1–27.9] | 0.70 [0.62–0.78] | 0.82 [0.73–0.93] |
| >20 | 22.8 [19.9–25.9] | 0.59 [0.49–0.70] | 0.69 [0.57–0.83] |
| Main type of cigarettes smoked[3] | | | |
| Manufactured | 31.1 [30.0–32.2] | Ref | Ref |
| Hand-rolled | 28.8 [27.7–29.9] | 0.90 [0.83–0.97] | 0.82 [0.75–0.88] |
| Strength of urges to smoke | | | |
| Not at all | 19.0 [17.5–20.5] | Ref | Ref |
| Slight | 28.2 [26.7–29.8] | 1.68 [1.48–1.91] | 1.77 [1.55–2.01] |
| Moderate | 31.2 [30.0–32.4] | 1.94 [1.73–2.17] | 2.12 [1.89–2.39] |
| Strong | 35.6 [33.6–37.5] | 2.36 [2.07–2.69] | 2.67 [2.33–3.06] |
| Very strong | 37.4 [34.2–40.8] | 2.55 [2.14–3.03] | 2.80 [1.69–2.58] |
| Extremely strong | 30.7 [27.0–34.6] | 1.89 [1.54–2.32] | 2.08 [1.69–2.58] |

CI, confidence interval. OR, odds ratio.

[1] Odds of having made a past-year smoking quit attempt, adjusted for survey year.

[2] Odds of having made a past-year smoking quit attempt, adjusted for survey year, age, gender, ethnicity, and social grade.

[3] Among those who currently smoke cigarettes.
